# INTRAVENOUS VITAMIN C SUPPLEMENTATION IN ALLOGENEIC HEMATOPOIETIC CELL TRANSPLANT RECIPIENTS: SALUTARY IMPACT ON CLINICAL OUTCOMES

**DOI:** 10.1101/2023.10.24.23297165

**Authors:** Gary Simmons, Roy Sabo, May Aziz, Erika Martin, Robyn J. Bernard, Manjari Sriparna, Cody McIntire, Elizabeth Krieger, Donald F. Brophy, Ramesh Natarajan, Alpha Fowler, Catherine H. Roberts, Amir Toor

**Author notes:** Denotes authors who contributed equally to this work (Manjari Sriparna, Cody McIntire). Correspondence, Amir A. Toor, MD,; Medical Director Stem Cell Transplant Program, Lehigh Valley Health Network, Allentown, PA and Affiliate Professor of Medicine, Department of Internal Medicine and Pediatrics, Virginia Commonwealth University, Richmond, VA.

## Abstract

Intravenous (IV) vitamin C improves organ function and reduces inflammation in sepsis, an inflammatory state like the post-hematopoietic stem cell transplant (SCT) milieu. The safety and efficacy of parenteral vitamin C after allogeneic hematopoietic stem cell transplant (HSCT) were evaluated in a phase I/II trial and clinical outcomes compared with a propensity score - matched historical control.

**Methods:** Patients with advanced hematologic malignancies were enrolled in a phase 2 clinical trial, receiving IV vitamin C, 50mg/kg/d, divided into 3 doses given on days 1-14 after HSCT, followed by 500 mg bid oral from day 15 until 6 months post-SCT.

**Results:** 55 patients received IV vitamin C: these include 10/10 HLA-MRD and MUD (n=48) and 9/10 HLA MUD recipients (n=7). All patients enrolled were deficient in vitamin C at day 0 and had restoration to normal levels for the remainder of the course. Vitamin C recipients had lower non-relapse mortality (11% vs. 25%, p-value = 0.07) and consequently, improved survival compared to historical controls (82% vs 62% p=0.06), with no attributable grade 3 and 4 toxicities to vitamin C. Patients with myeloid malignancies had improved survival (83% vs. 54%, p=0.02) and non-relapse mortality (NRM) (10% vs. 37%, p=0.009), as well as chronic GVHD, with similar relapse rates compared to controls.

**Conclusions:** In patients undergoing allogeneic HSCT the administration of IV vitamin C is safe and reduces non-relapse mortality improving overall survival. Randomized trials are needed to confirm the utility of this easily available and inexpensive therapy.

## INTRODUCTION

Hematopoietic stem cell transplant (HCT) is a potentially curative therapy provided by the high dose intensity of the preparative regimens used and the immune mediated graft vs leukemia effect^1^. Preparative regimens such as myeloablative conditioning with ionizing radiation and alkylating agents like busulfan or melphalan, work in large part by widespread, indiscriminate oxidative damage to DNA. Consequently, these oxidative stress produced by these regimens induces variable degree of enteral mucositis, loss of functioning epithelial cells and disrupt tight junctions increasing the translocation of bacterial products and inflammatory cytokines, which is considered an essential factor in the pathogenesis of acute graft vs host disease GVHD.^2^ This tissue injury leads to endogenous tissue antigen presentation to donor T cells provoking GVHD,^3^ contributing to 1-year transplant related mortality (TRM) in myeloablative allogeneic HCT, which has been reported between 15-31 %.^4^ ^5^

It is critical that therapy to mitigate tissue injury in HSCT recipients be developed to aid in reducing GVHD incidence contributing to TRM. An antioxidant agent that may serve this purpose is the micronutrient ascorbic acid, vitamin C. Aside from its anti-oxidant effect, It is an active anti-inflammatory agent with the ability to inhibit NF-κB-driven inflammatory cytokine (IL-6, IL-8, and TNF-α) expression and to attenuate endothelial permeability.^6^ ^7^ Hypovitaminosis C is well described in critically ill patients, who have several parallels with HSCT patients, such as, elevation of biomarkers including C-reactive protein (CRP), as well as having elevated levels of inflammatory cytokines. In sepsis patients treated with parenteral vitamin C, organ function improves, and inflammatory markers decline.^8^

In previous reports, vitamin C deficiency was observed during the acute phase of HCT and this was significantly associated with elevated levels of inflammatory markers, CRP, and ferritin.^9^ Vitamin C administered in enteral nutrition may not be adequate due to poor absorption and increased tissue utilization.^10^ Previous work in a prospectively studied cohort of patients undergoing myeloablative conditioning allogeneic HSCT, found patients to be uniformly vitamin C deficient and these individuals remained deficient up to 60 days post-transplant. Patients with severe mucositis had lower vitamin C levels at day 14 after HSCT.^11^

Recognizing that vitamin C has anti-inflammatory properties and patients undergoing allogeneic HSCT are deficient in this critical micronutrient, likely due to increased consumption and reduced dietary intake and absorption, a phase II study was developed to prospectively administer parenteral vitamin C following transplantation. This was done to study the hypothesis that mitigating the pro-inflammatory effects of HCT with early administration of vitamin C in patients deficient in this micronutrient will attenuate endothelial and organ injury from high-dose conditioning and ameliorate GVHD and transplant-related mortality. The results of this trial are now reported, comparing patients who received parenteral vitamin C after myeloablative conditioning for allogeneic HSCT to propensity-matched historical controls.

## METHODS

### Patients

This was a prospective phase II clinical trial with a safety lead-in cohort, designed to study the impact of parenteral vitamin C administration in recipients of allogeneic SCT (FDA_IND 138924). This trial was approved by the Massey Cancer Center Protocol Review Committee and Human Subjects Institutional Review Board (IRB) at Virginia Commonwealth University (VCU). All patients signed IRB-approved informed consent forms and were treated at VCU (NCT03613727). The study population included adult patients 18-78 years old with acute myelogenous leukemia (AML), acute lymphoblastic leukemia (ALL), chronic myelogenous leukemia (CML), and myelodysplastic syndrome (MDS) who underwent their first allogeneic stem cell transplant from matched sibling donors and unrelated donors who were matched at either 7/8 or 8/8 HLA-A, -B, -C, -DRB1 loci using high-resolution DNA-based typing. Patients undergoing non-myeloablative conditioning were not included.

### Vitamin C administration

An initial cohort of patients (N=14) were enrolled if they had low vitamin C levels (<0.5mg/dl). Vitamin C levels were checked at baseline and on day −2 in these patients. Patients were treated with parenteral vitamin C 50 mg/kg/d (Ascorbic Acid; McGuff Pharmaceuticals, Santa Ana, CA) divided in three doses from day +1 through day +14 and transitioned to oral vitamin C 500 mg twice daily from day +15 through day +180 (**Supplementary Figure 1**). Vitamin C was given in 50 mL of 5% dextrose and water over 30 minutes every 8 hours in UV-light-protected bags and tubing. Subsequently consecutive patients meeting inclusion criteria were enrolled in the trial till study completion (N=55).

### Cytokine analysis

Vitamin C and CRP were checked before conditioning (baseline) and on the following days: day 0, 14, 30, 100 in all the patients. In the first 14 patients enrolled, pro-inflammatory cytokines IL-1b, IL-2, IL-6, IL-12, TNF-α, IFN-γ, as well as soluble thrombomodulin were measured at baseline and on days 0, 14 and 30; IL4, IL-5, IL-10, IL-13, IL-17A/CTLA8, IL-21, IL-22, IL-23, were also measured at these times.

Cytokines were assayed in patient plasma with multiplex technology using the xMAP Luminex 200 (Luminex Corp, Austin TX). These data were displayed in heat maps where, to normalize the cytokine measurements, the cytokine levels at each time point in each patient (days 0, 14, 30, 100) were divided by the corresponding baseline measurements for that patient. Further grouping of the resulting data was completed via conditional formatting in MS Excel into low (green), medium (yellow) and high (red) valued cytokine levels to give a visual representation of change over time. CMV reactivation was defined as > 200 IU/mL; EBV reactivation as a titer > 500 IU/mL and requiring Rituximab.

### Statistical Analysis

The primary endpoint was non-relapse mortality at one year and secondary endpoints included engraftment, acute and chronic GVHD, relapse, and overall survival. Simon’s two-stage design was utilized for this study. In the first stage, a threshold of ≥35% NRM was set to result in trial termination for futility. The total sample size calculated for Simon’s two-stage design was 55 patients for a reduction in NRM from 35% to 20%, a 42% relative reduction compared to a propensity-matched historical control cohort. Propensity score matching was performed to identify similarly treated patients from a historical control cohort, transplanted between January 2015 to December 2018. To accomplish this, a logistic regression was fit with the intervention group as the binary outcome (intervention vs. historical control), against three matching variables, including diagnosis (ALL, AML, and CML+MDS), conditioning regimen (busulfan + cyclophosphamide, fludarabine + melphalan, and total body irradiation+cyclophosphamide), and CIBMTR disease risk index (high and intermediate+low). The resulting probabilities of belonging to the Intervention group were used as propensity scores. Nearest neighbor matching was used to identify the matching set of historical control subjects in a 1:1 ratio with the trial enrollees.

A Kaplan-Meier step function was performed for mortality assessment, while cumulative incidence curves were constructed for relapse (accounting for competing risk of mortality), acute graft versus host disease (GVHD), chronic GVHD (accounting for competing risks of relapse and mortality), non-relapse mortality (NRM). Cox-proportional hazard models were used to estimate adjusted hazard ratios between the time-to-event outcomes and group, adjusted for patient age, donor type, stem cell source, disease type, conditioning regimen, and disease risk. A subset analysis was performed analyzing outcomes in patients with AML, MDS and CML. The R (4.1.2) and RStudio (version 2022.02.0) statistical software platforms were used for all data management and analysis.

### Clinical outcome graphs

To examine the relative impact of variables influencing clinical outcomes, graphs depicting clinical outcomes were utilized. Graphs are mathematical constructs in which nodes, or vertices are connected with edges which may be unidirectional or bidirectional. This form of network analysis allows easy visualization of relationships between the entities occupying the various nodes with the intervening edges (**Supplementary Figure 2**). For this analysis the transplant event, clinical states (acute and chronic GVHD, relapse) and outcomes (TRM and OS) were modeled as vertices, and the edges connecting them were considered to have been weighted by the variables examined in the multivariate analyses. Since these were calculated for each transitional state and final outcome in the total cohort, these measures are depicted at the nodes/vertices, but in effect their magnitude represents the probability of an individual taking a path along a particular edge to a given node (or clinical state/outcome). The graph methodology provides an easily visualizable means of assessing the relative effect of different variables on clinical outcomes in HCT where multiple inter-related clinical outcomes are studied, potentially simplifying complex data interpretation.

## RESULTS

### Vitamin C following allogeneic SCT

Patients were prospectively enrolled in this study between October 2018 – October 2021 and propensity scores matched with historical controls transplanted between 2015 – 2018. Follow up was updated as of February 2023. Patient characteristics are described in **Table 1**; 55 patients received IV vitamin C, including 10/10 HLA-MRD and MUD (n=48) and 9/10 HLA MUD recipients (n=7; 4 HLA-DQ, 2 HLA-B and 1 HLA-A mismatch). The number of fludarabine and melphalan recipients was higher in the vitamin C group as compared to historical controls.

**TABLE 1:**
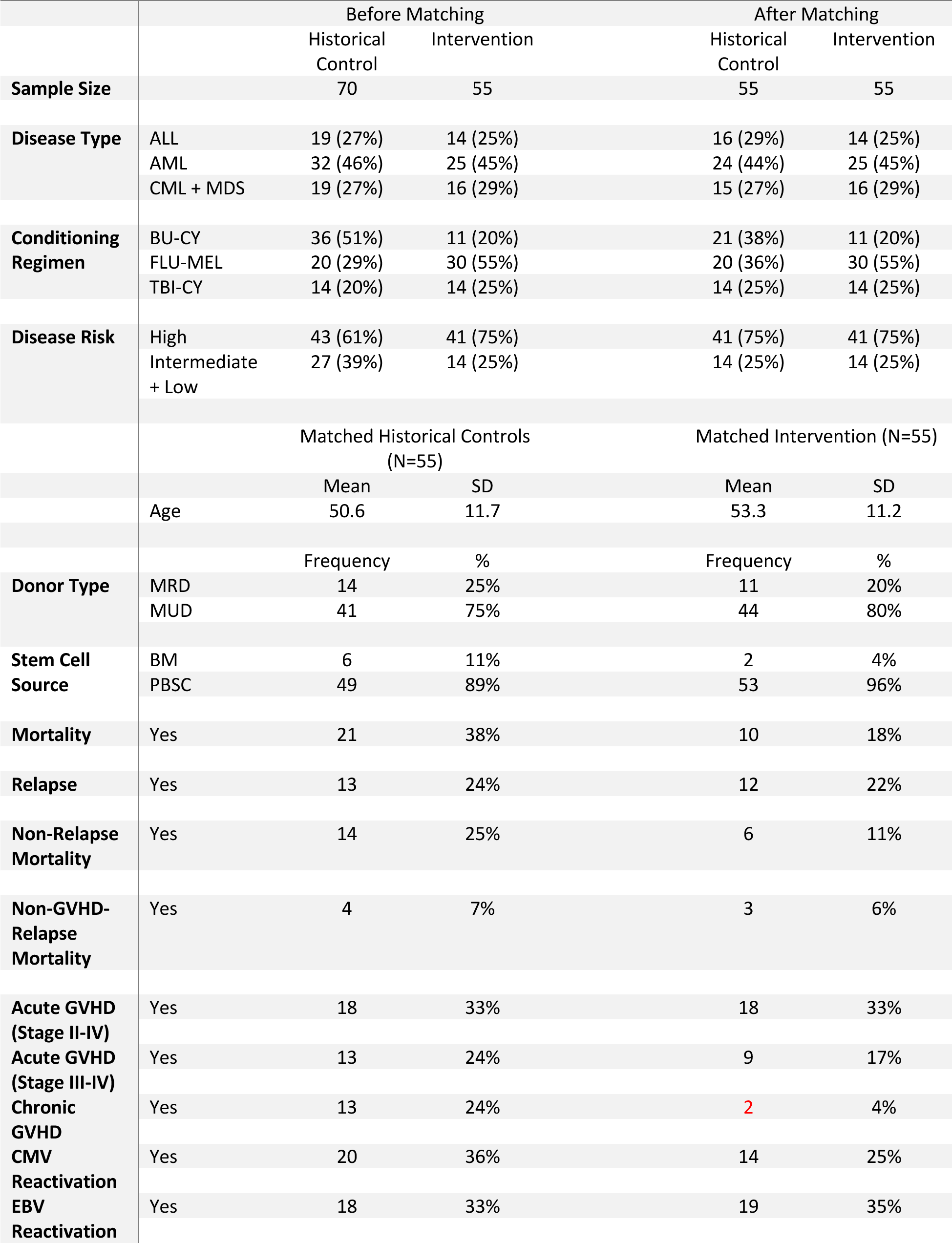
Summaries for matching variables and other patient measurements (before and after matching)

A safety lead-in cohort of 14 patients was initially enrolled. All patients enrolled were deficient in vitamin C at day 0, with a median level of 0.3 mg/dL (range: 0.1-0.5); the median neutrophil recovery was by 12 days (range 9-15 days) in these patients; platelets engrafted by day 12 (8-21 days), and 100% donor myeloid engraftment was recorded at day 30. Safety endpoints of lack or loss of myeloid engraftment, grade 3 acute GVHD and NRM were evaluated in these patients, without triggering any of the stopping thresholds.

### Engraftment in vitamin C recipients

In the entire cohort of patients treated with Vitamin C (N=55), the median times to neutrophil and platelet engraftment were 11 and 12 days respectively (range, neutrophils: 9-15 days, platelet 8-21 days). T cell chimerism at day 30, 60, and 90 following HSCT was > 95% in the Vitamin C group. Concomitantly measured median CD3+ cell count on days 30, 60, and 90 were 236 (range: 2-2024), 485 (5-5580) and 390 (112-2565)/μL respectively and were not different compared with the historical controls. Median CD3+/4+ T cell count on day 30 was 80/μL (range 30-416), CD3+/8+ T cell count was 198/μL (25-1924), CD19+ cell count was 8/μL (range 5-84) and CD3-/56+ NK cell count was 260/μL (range 25-656), indicating early T and NK cell recovery in these patients.

### Clinical outcomes in vitamin C recipients

Clinical outcomes were generally improved in vitamin C recipients. Graph analysis demonstrated that there was a trend for greater likelihood of acute and chronic GVHD, as well as TRM in MUD recipients, leading to a significantly lower survival in these cohorts (**Figure 1**). HCT recipients with AML had a greater likelihood of relapse and subsequently, a significantly lower likelihood of survival. Vitamin C had a salutary impact on all these outcomes, with a trend for lower non-relapse mortality in the vitamin C treated group (11%) compared to the historical control (25%) (HR = 0.4, 95% CI: 0.1, 1.0, p-value = 0.069) (**Figure 2**). Overall survival was also improved in study participants ((HR = 0.5, 95% CI: 0.2, 1.0, p-value = 0.065) (**Figure 3**).

**Figure 1.**
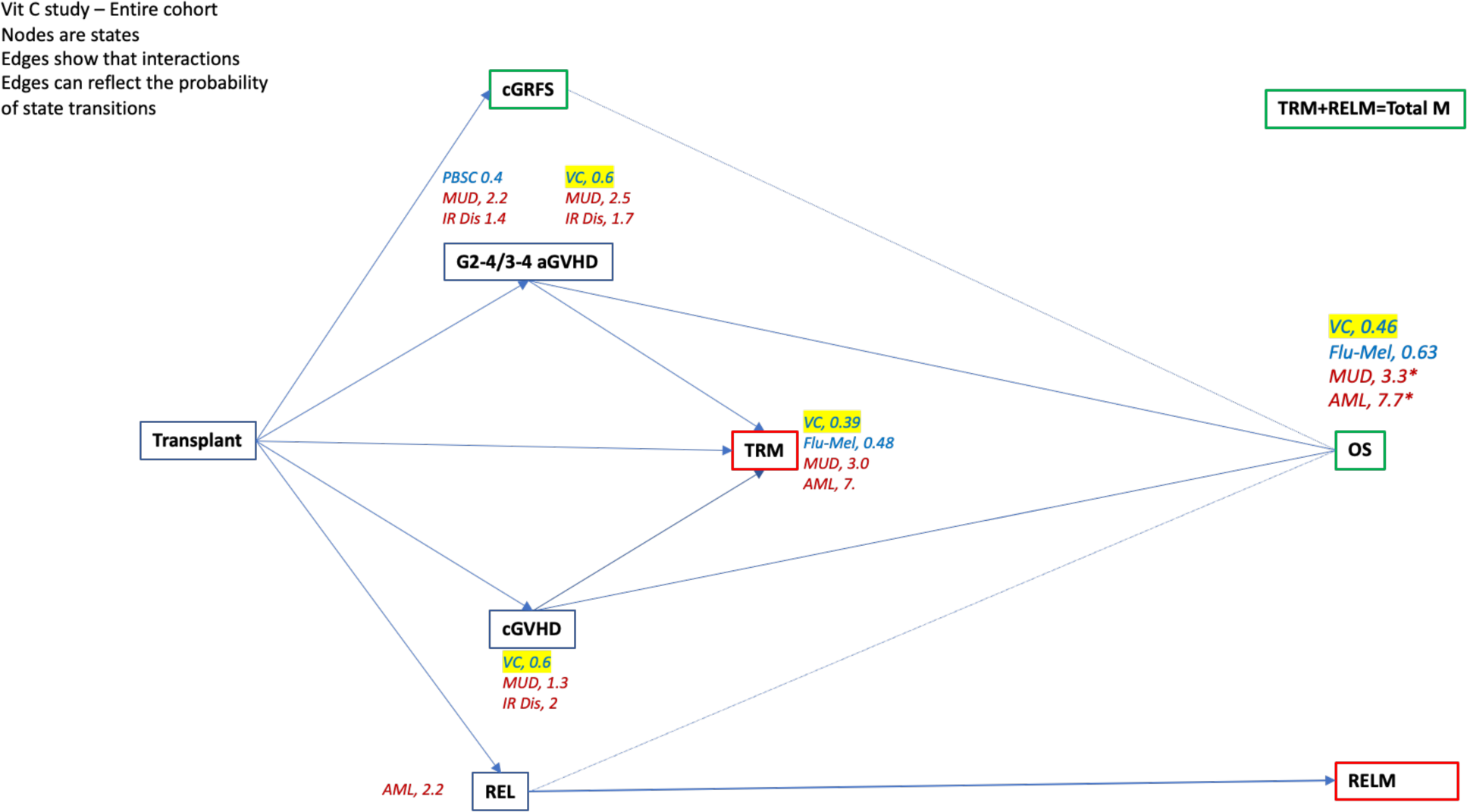
Graph depicting sequential outcomes in vitamin C recipients and historical controls following transplant. First tier clinical outcomes include GVHD and relapse, transient states which lead to clinical outcomes, survival, and mortality depicted in second tier nodes.

**Figure 2.**
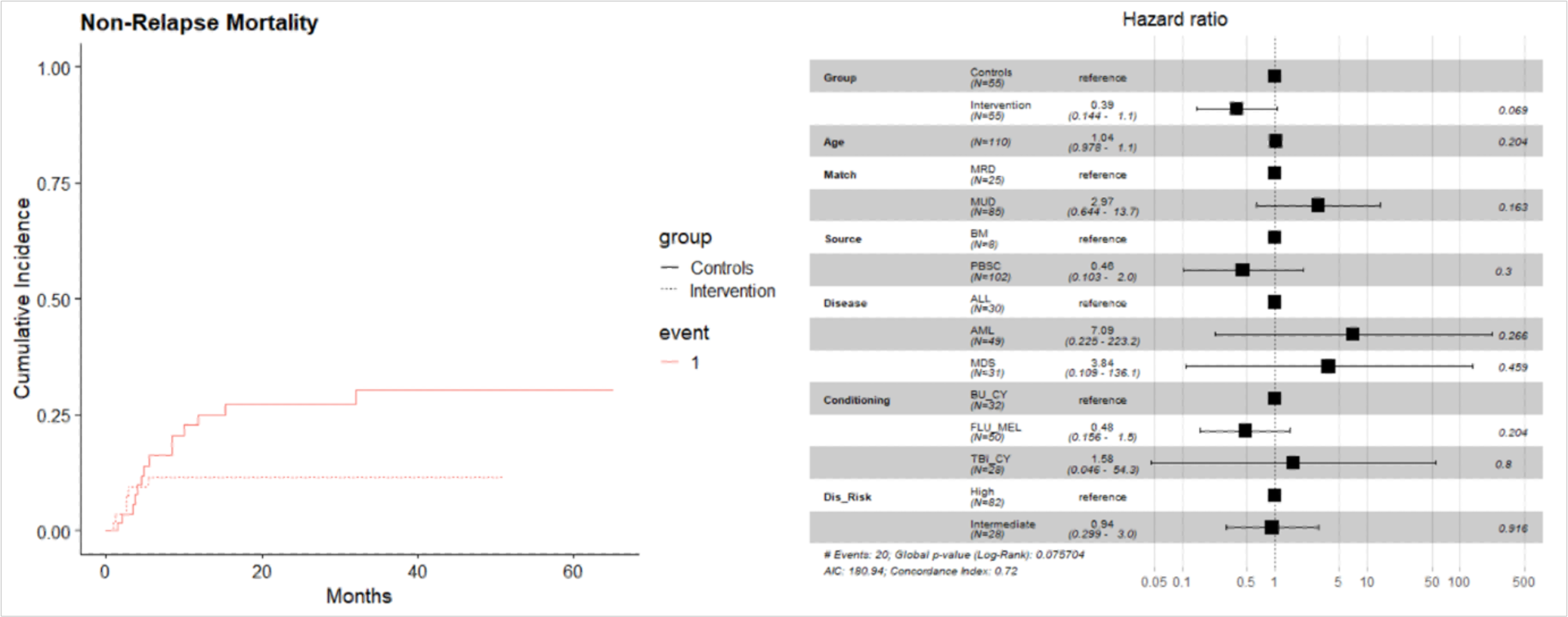
(A) CI curves depicting lower NRM in the VC treated patient. (B) In a multivariate model this was not significant (HR = 0.4, 95% CI: 0.1, 1.0, p-value = 0.069).

**Figure 3.**
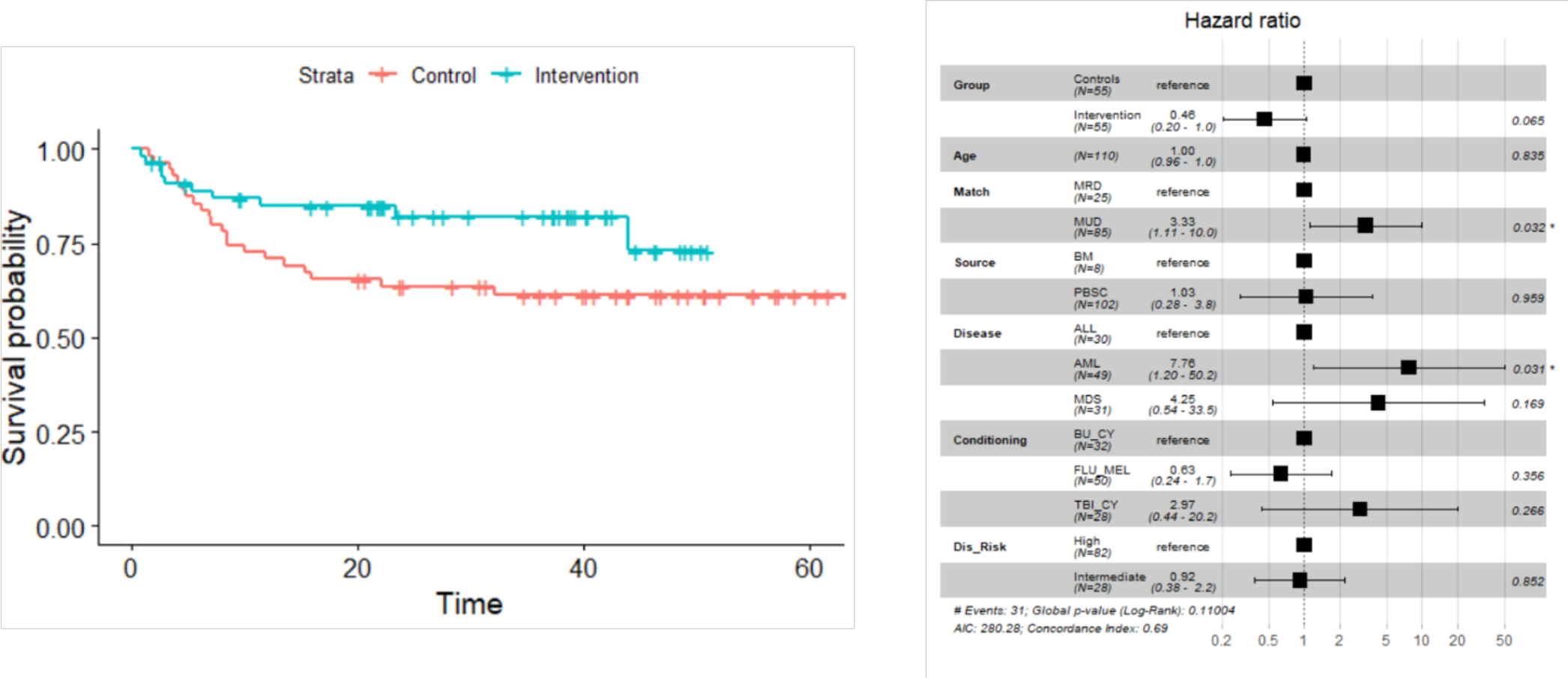
(A) KM curves depicting improved survival in the VC treated patients (p= 0.057). (B) In a multivariate model, only donor type (MUD vs. MRD: HR = 3.3, 95% CI: 1.1, 10.0, p = 0.032) and diagnosis (AML vs. ALL: HR = 7.75, 95% CI: 1.2, 50.2, p = 0.031) were significant.

The vitamin C cohort had 7/55 HLA mismatched unrelated donor HCT recipients, despite this there was no increase observed in the risk of acute GVHD in the control vs. study cohorts (grade II-IV, (33%) vs. (33%), p-value = 0.81 or III-IV, (24%) vs. (17%), p-value = 0.35). Organ distribution of acute GVHD grade 3-4 in vitamin C recipients was; skin only (2), GI (3), skin and liver (1), skin and GI (2) and steroid refractory GI (2). Moderate to severe chronic GVHD rates when accounting for competing risks of relapse and mortality were lower in the Vitamin C group (11%) compared to the historical controls (24%), (adj HR = 0.60, 95% CI: 0.20, 1.76, p-value = 0.352) (**Supplementary Figure 3**).

Patients in the vitamin C vs. historical control cohorts received rabbit anti-thymocyte globulin (ATG) as a part of GVHD prophylaxis regimen. Of note, the schedule of ATG administration was predominantly day −3 to −1 in the historical control and day −9 to −7 in the Vitamin C recipients, with similar rates of acute and chronic GVHD observed despite the earlier administration of ATG, and a lower *in vivo* T cell depleting effect.

### Clinical outcomes in myeloid malignancy

Because of the frequency of DNA hypermethylation in myeloid malignancies, the clinical outcomes in patients with AML, MDS and CML were examined comparing trial patients with their propensity-matched historical controls (**Table 2**). Graph analysis again demonstrated the adverse impact of MUD and AML on sequential outcomes, but in this myeloid malignancy cohort the trend for a salutary impact of vitamin C on clinical outcomes, extended beyond acute and chronic GVHD to disease relapse, with a consequent statistically significant benefit observed in survival (**Figure 4**). Overall survival was superior amongst the vitamin C recipients with myeloid malignancy (p = 0.0167) (HR = 0.32, 95% CI: 0.12, 0.84, p = 0.02) (**Figure 5**), in large part attributable to a lower risk of NRM in these patients (10%) compared with historical controls (37%) (HR = 0.22, 95% CI: 0.07, 0.69, p-value = 0.009) (**Figure 6**). The rate of chronic GVHD was lower in Vitamin C recipients (12 vs. 24%; adj HR = 0.56, 95% CI: 0.17, 1.82, p-value = 0.383) between the two groups.

**Figure 4.**
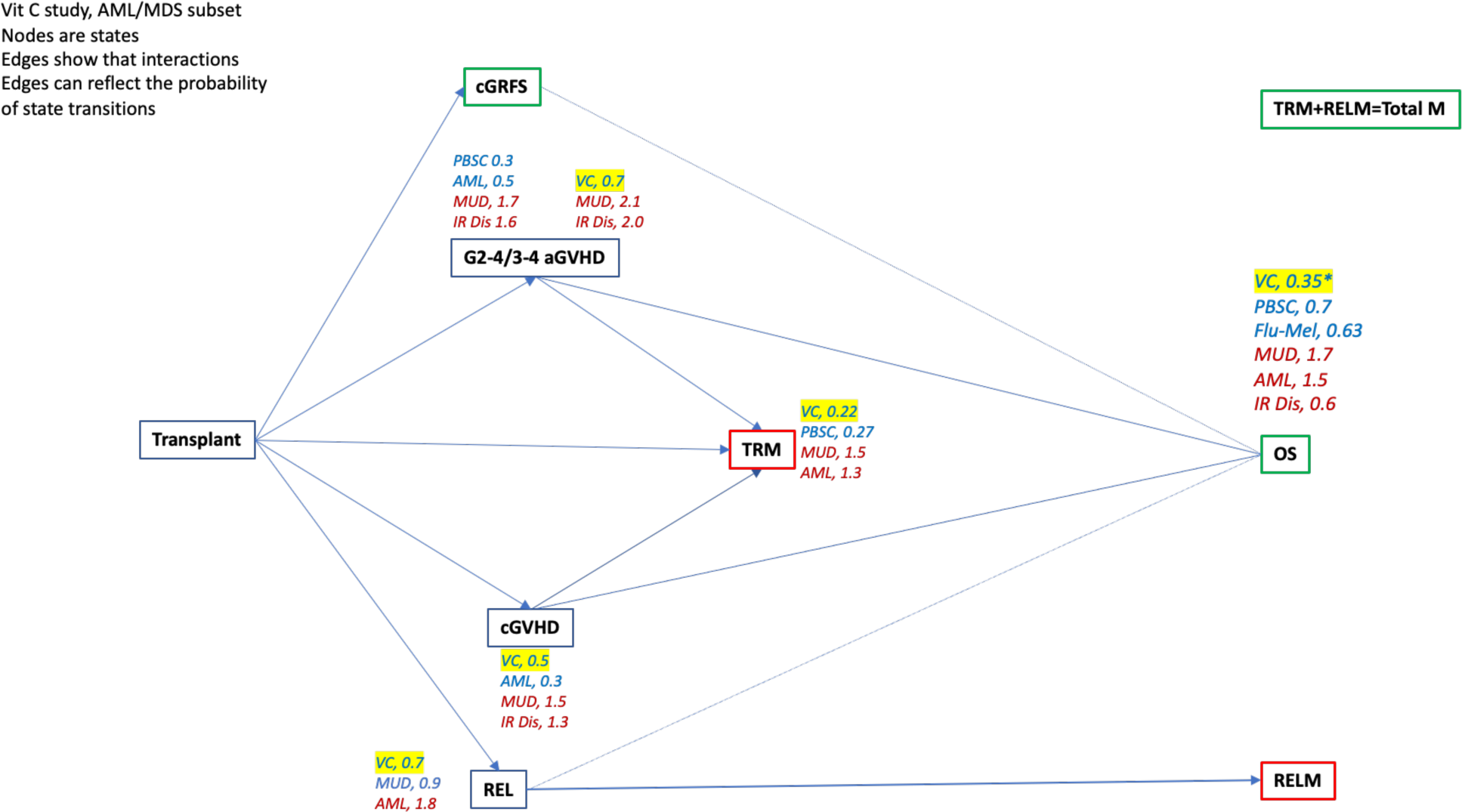
Graph depicting sequential outcomes in vitamin C recipients and historical controls with myeloid malignancy following transplant. First tier clinical outcomes include GVHD and relapse, transient states which lead to clinical outcomes, survival, and mortality depicted in second tier nodes.

**Figure 5.**
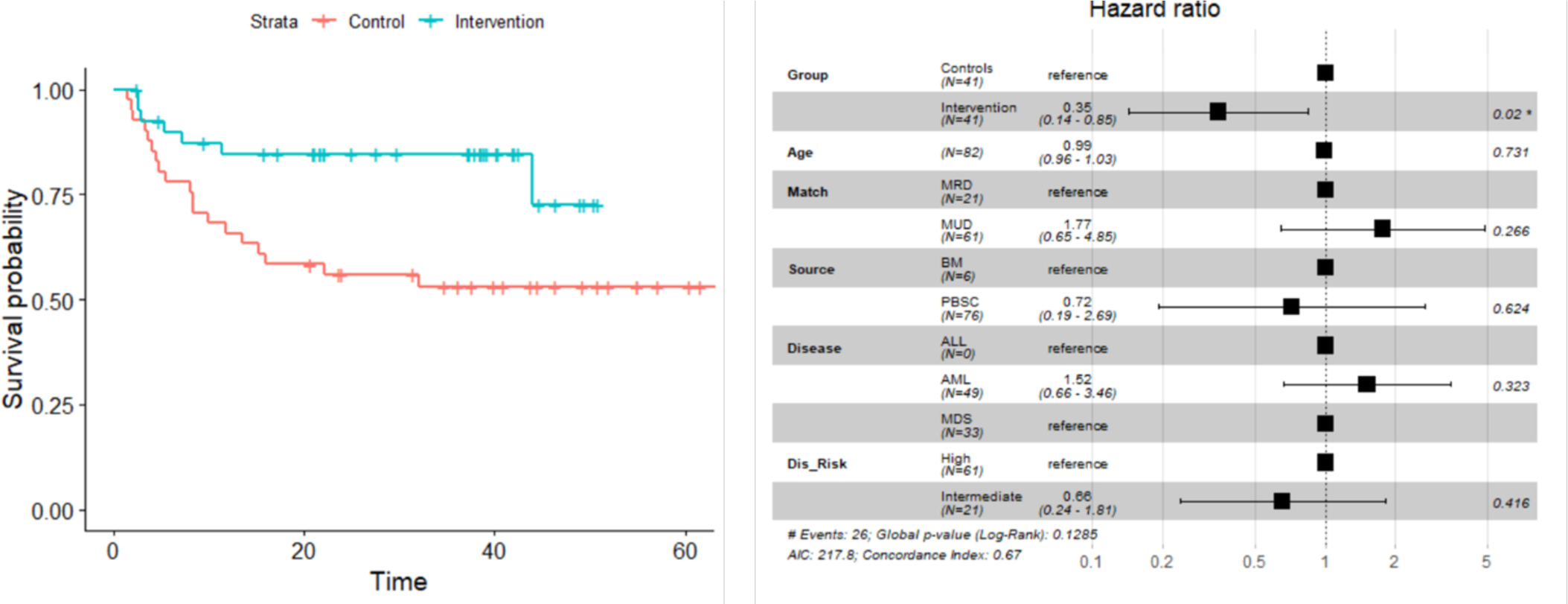
(A) KM curves depicting improved survival in the VC treated patients with myeloid malignancies (p= 0.0167). (B) In a multivariate model this relationship remained significant (HR = 0.32, 95% CI: 0.12, 0.84, p-value = 0.02) amongst myeloid malignancy patient.

**Figure 6.**
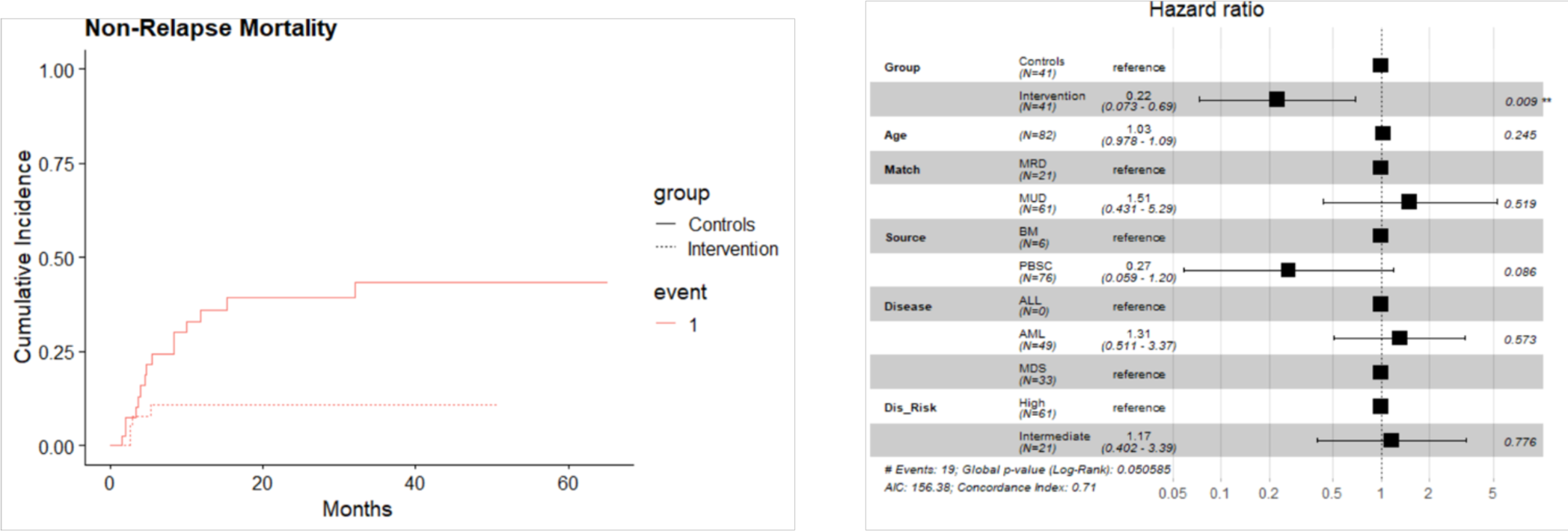
(A) CI curves depicting lower NRM in the VC treated patients with myeloid malignancies. (B) In a multivariate model this was significant (HR = 0.22, 95% CI: 0.07, 0.69, p-value = 0.009).

**TABLE 2:**
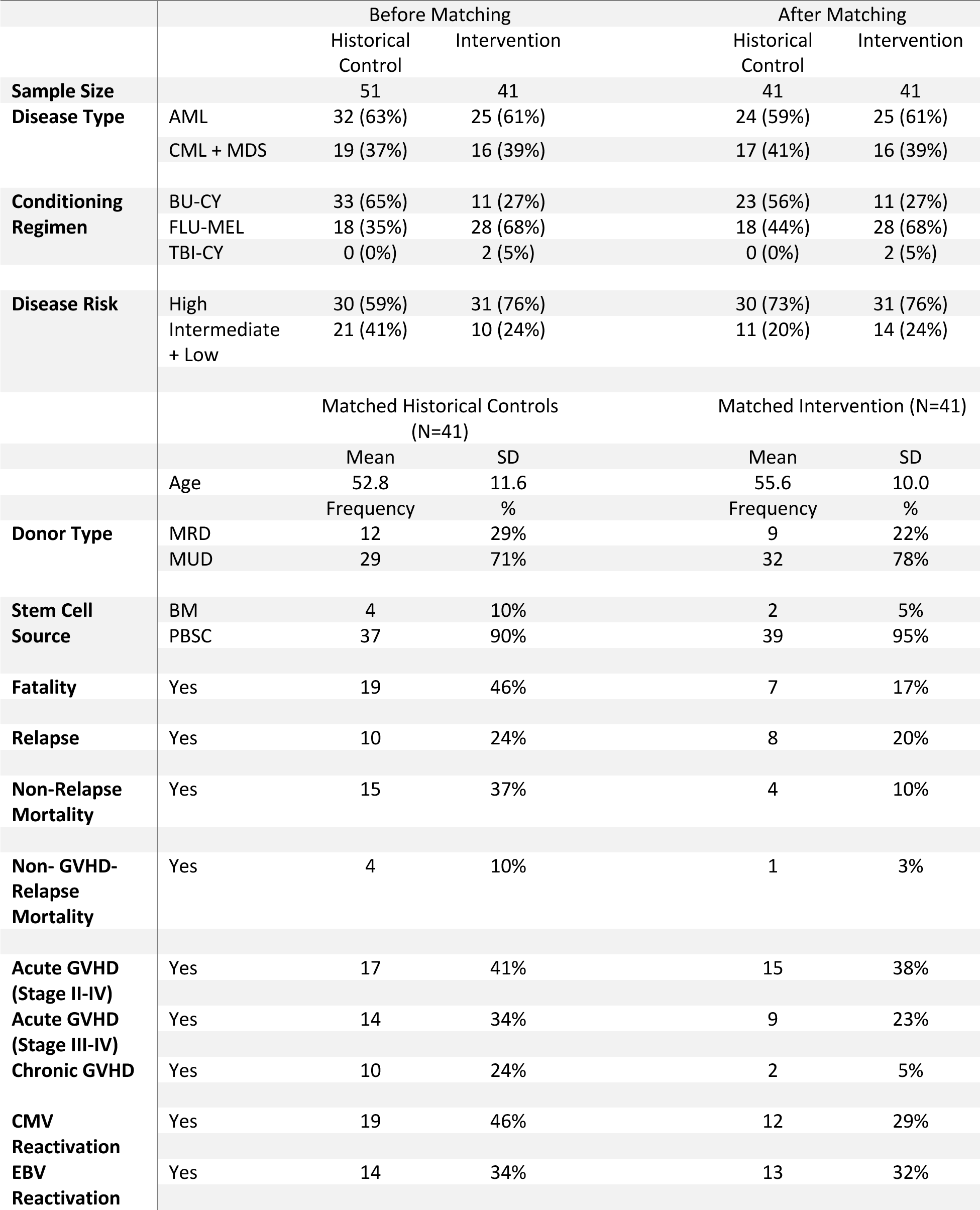
Summaries for matching variables and other patient measurements (before and after matching)

### Infections and toxicities in the Vitamin C recipients

CMV reactivation rate was higher in historical controls (36%) than intervention (24%) (p-value = 0.35), and a similar trend was observed in myeloid malignancy patients. EBV reactivation rate was similar in historical controls (33%) and vitamin C recipients (35%) (p-value = 0.88). There were no grade 3 – 5 adverse events attributable to vitamin C therapy in this trial, particularly no nephrotoxicity or renal calculi were observed.

### Cytokine levels in Vitamin C recipients

Vitamin C level (normal 0.4-2.0 mg/dL) was universally low prior to intervention and was restored to normal by day 14 (**Figure 7**), while CRP (normal 0-0.5 mg/dL) rose in the early aftermath of transplant following conditioning and engraftment, and came down later. The peaks for both the parameters coincided, suggesting potential benefit of vitamin C may have come from its physiologic effect at the time of maximum inflammatory cytokine surge. The first 14 patients treated with vitamin C had pro-inflammatory cytokines IL-1b, IL-2, IL-6, IL-12, TNF-α, IFN-γ, and soluble thrombomodulin measured, which remained unchanged between day 0 and day 14, as well as day 30 after HCT (P=NS for all direct comparisons). Heat maps demonstrating change of cytokine levels post-transplant showed that most of the patients showed either a reduction or stabilization of their inflammatory cytokine levels following SCT, with few patients demonstrating an increase at all the time points tested following HSCT (**Supplementary Figure 4**). However, in most instances the cytokine levels were back down in the lower ranges by day 30. Elevated cytokine levels on day 14 appear to correspond to higher magnitude of donor derived T cell recovery by day 30 following transplant (**Supplementary Figure 5**).

**Figure 7.**
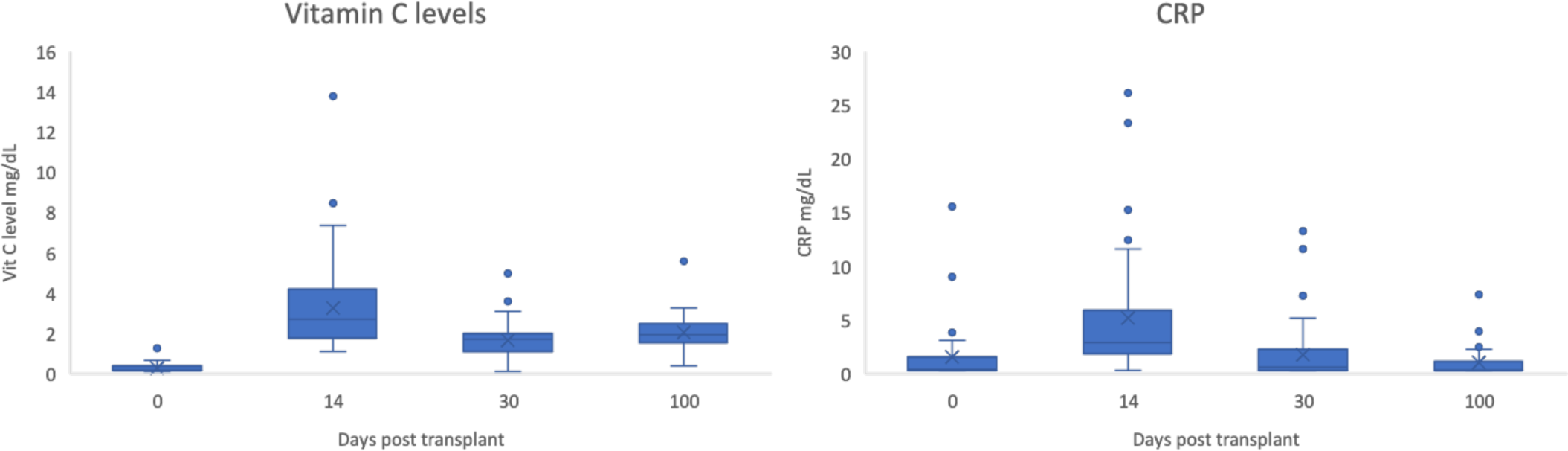
(A) Vitamin C levels in the entire cohort of patients. (B) CRP levels at the same time points.

## DISCUSSION

In this paper the results of a prospective trial evaluating parenteral vitamin C in allogeneic HCT are presented, demonstrating its safety. There was a non-relapse mortality and survival benefit observed, especially in recipients with myeloid malignancies when compared with propensity-matched historical controls, adjusting for diagnosis, disease risk, and conditioning intensity. Graph analysis of sequential clinical outcomes indicates possible impact of vitamin C on GVHD risk may have impacted these two clinical outcomes.

Non-relapse mortality is the Achilles heel of allografting, compromising the benefit that comes from the immune graft vs. malignancy effect. The foundation for NRM is laid in the initial mucosal and endothelial injury seen following high-dose therapy, which sets the stage for alloantigen presentation to donor cells, setting up the cascade of alloimmune response culminating in GVHD.^12, 13^ Pro-inflammatory cytokines released following tissue injury from high-dose therapy is a major contributor to the pathophysiology in these instances.^14^ This is similar to sepsis, where parenteral vitamin C has been shown to ameliorate endothelial injury, especially in lung injury and acute respiratory distress syndrome. ^8^ Further due to the oxidative stress of pretransplant conditioning with radiation and alkylating agents,^15^ ^16^ followed by poor oral intake, the vitamin C stores are rapidly depleted,^17^ leading to a severely deficient state. ^11^ ^18^ ^19^ Given this logic the clinical trial reported here was designed to rapidly restore vitamin C following transplantation and utilize its endothelial stabilizing effect to reduce the impact of mucosal and endothelial injury post-transplant.^20^ ^21^ Ameliorating the tissue injury in turn would modulate the inflammatory milieu, potentially mitigating alloantigen presentation in the early days following transplant, eventually reducing GVHD likelihood evolving from recovering donor derived T cells. This concept is supported here by the observed trend toward reduced acute GVHD (grade II-IV) and chronic GVHD in Vitamin C recipients, as visualized in the graph analysis. The reduction in chronic GVHD incidence was observed in vitamin C recipients compared to historical controls despite an equivalent number of HLA MUD and PBSC recipients, and a different schedule of ATG administration designed to enhance donor T cell recovery.^22^ Lower GVHD risk was followed by a lower risk for NRM and downstream benefit in survival for patients in the intervention cohort, particularly in those with myeloid malignancy. In these patients, despite the reduction in chronic GVHD, relapse risk was diminished, even though a higher proportion of vitamin C recipients underwent conditioning with fludarabine and melphalan, a reduced intensity regimen. This points to a potential modulation of disease risk impact in addition to attenuating GVHD in these high-risk leukemia patients.

Vitamin C is a cofactor for the TET-2 enzyme, and may have a role in reducing relapse risk in patients with mutation impacting this mechanism of hypomethylation.^23^ Vitamin C by helping restore the oxidative function of TET-2 would reduce the risk of relapse in myeloid malignancies. ^24^ ^25^ ^26^ ^27^ While the relapse rate is not significantly different in this cohort, those relapsing had high-risk disease, and included patients who were transplanted with active disease and primary induction failure. A further confounding factor was that most patients underwent GVHD prophylaxis with ATG, potentially limiting the GVL effects that may be observed. One may surmise that vitamin C mediated relapse protection effect may be most evident in patients who do not get in vivo T cell depletion. This will also be the population where any GVHD benefit from diminished alloantigen presentation may be best observed.

A reduction in inflammation may increase the risk of infections. Infections were seen in the vitamin C recipients, but not at a higher rate than the matched historical controls, nevertheless given a reduction in the inflammatory response, this bears close observation in future trials, as was suggested by a randomized trial of vitamin C administration in patients with early sepsis.^28^ Such a trial, of necessity, must be randomized, and optimally conducted in patients who do not receive T cell depletion (in vivo or ex vivo). Parenteral vitamin C has been reported to lead to an improvement in biomarker profile in sepsis, consistent with that, this study also demonstrated an amelioration of adverse cytokine profiles following high-dose therapy and HSCT.

If vitamin C repletion following high-dose therapy and HSCT is proven to be of benefit in patients undergoing T cell replete allografts and reduce relapse risk, it will be a significant advance on a global scale, particularly in economically challenged regions, given its low cost and wide availability. Therefore, our study demonstrating the absence of increased toxicity and impact on GVHD, relapse and non-relapse mortality represents a crucial first step in that direction.

## Data Availability

All data produced in the present study are available upon reasonable request to the authors

## Acknowledgements

This trial was supported by research funding from the NIH-NCI Cancer Center Support Grant (P30-CA016059; PI: Gordon Ginder, MD).

## Author contributions

Gary Simmons: Develop and conduct study, collect and analyze data, write paper; Roy Sabo: Develop and conduct study, analyze data, write paper; May Aziz: Conduct study; Erika Martin: Conduct study; Robyn Bernard: Collect data; Manjari Sriparna: Collect data, write paper; Cody McIntire: Collect data, write paper; Elizabeth Krieger; review and write paper; Donald Brophy: Develop and conduct study, write paper; Ramesh Natarajan: Develop study, write paper; Alpha Fowler III: Conceptualize and develop study, write paper; Catherine H. Roberts: Develop and conduct study, collect and analyze data; and Amir Toor; Conceptualize, develop, conduct study, collect and analyze data.

## Supplemental Materials

Myeloablative conditioning included either total body irradiation (TBI) 12 Gray given from day −6 to day - 4 and cyclophosphamide 60 mg/kg/dose x 2 doses on days −3 and −2. Busulfan 0.8 mg/kg/dose x 16 doses from day −8 to day −5, with a target AUC of 5000 µMol*min/day, and cyclophosphamide 60 mg/kg/dose x 2 doses on day −3 and day −2. Fludarabine 30 mg/m2/dose x 4 doses on day −6 to day −3 and melphalan 140 mg/m^2^ on day −2.

GVHD prophylaxis included rabbit anti-thymocyte globulin (ATG) infused at a total dose of 5 mg/kg for unrelated donors and 3 mg/kg for related donors. Rabbit ATG was given over divided doses on day −3 to day −1 for patients receiving conditioning with TBI and cyclophosphamide or busulfan and cyclophosphamide and was given over day −9 to day −7 for patients receiving fludarabine and melphalan. All patients received a calcineurin inhibitor (cyclosporine or tacrolimus) and either short-course methotrexate 5-15 mg/m2/dose on day +1, +3, +6, and +11 (with leucovorin rescue) or mycophenolate mofetil 30 mg/kg/day in two divided doses on day +1 to day +30. Patients received both peripheral blood and bone marrow grafts. Engraftment was defined as absolute neutrophil count (ANC) > 500 x 3 days.

## Appendix: On Evaluating Cytokine Profiles

The dynamical systems theory of T cell biology allows one to consider the effects of the cytokine-cytokine receptor interactions on T cells. The cytokine mediated signal may be considered consisting of a matrix with cytokines and cytokine receptor vectors, because the cytokines and their receptors, have different magnitudes and varying receptor specific effects (directionality) on T cell growth and differentiation. Ignoring the di- or trimerization of cytokine receptor protein subunits, a simplified version of the cytokine tensor may be constructed as follows,

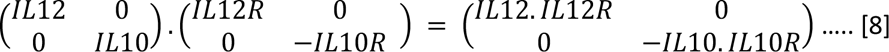

This is the cytokine vector, *Ck*, with the example showing the interaction between IL-12 and IL-10 and their respective receptors. It should be noted that cytokines may bind related receptors with different affinities, providing different vector components. The negative sign means a growth suppressive effect, the net effect of cytokines can either be negative or positive, the *Ck* can alter the magnitude and direction of T cell growth. Simplifying the above notation, we may consider that, each of the T cells or NK cells (any cell effected by cytokines for that matter), will then constitute a vector, and the cytokine milieu an operator.

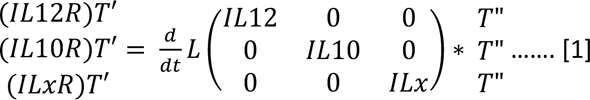

In this instance the T cell, in an initial state T”, has been transformed by the cytokine operator to a subsequent state T’, which may be an activated state or inactive state depending on the balance of cytokines, and its own cytokine receptor profile. The overall cytokine profile at any given time, t, will then, based on the T or NK cell array it encounters, and depending on the cytokine receptors being expressed by these cells, alter the immune profile of a post-transplant reconstituting immune system.

We have cytokine data but do not have cytokine receptor data. So, to glean information from this one might review the characteristics of the cytokine operator at different times and see how that correlates with clinical outcomes. To do so, one may simplify things even more by condensing the above identity matrix in equation to a column matrix

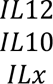

This done one may simply ‘cluster’ patients with similar cytokine profiles, create a heat map for easier visualization, and see if there is an association of specific clinical outcomes with different cytokine profiles.

---

**Supplementary Figure 1.**
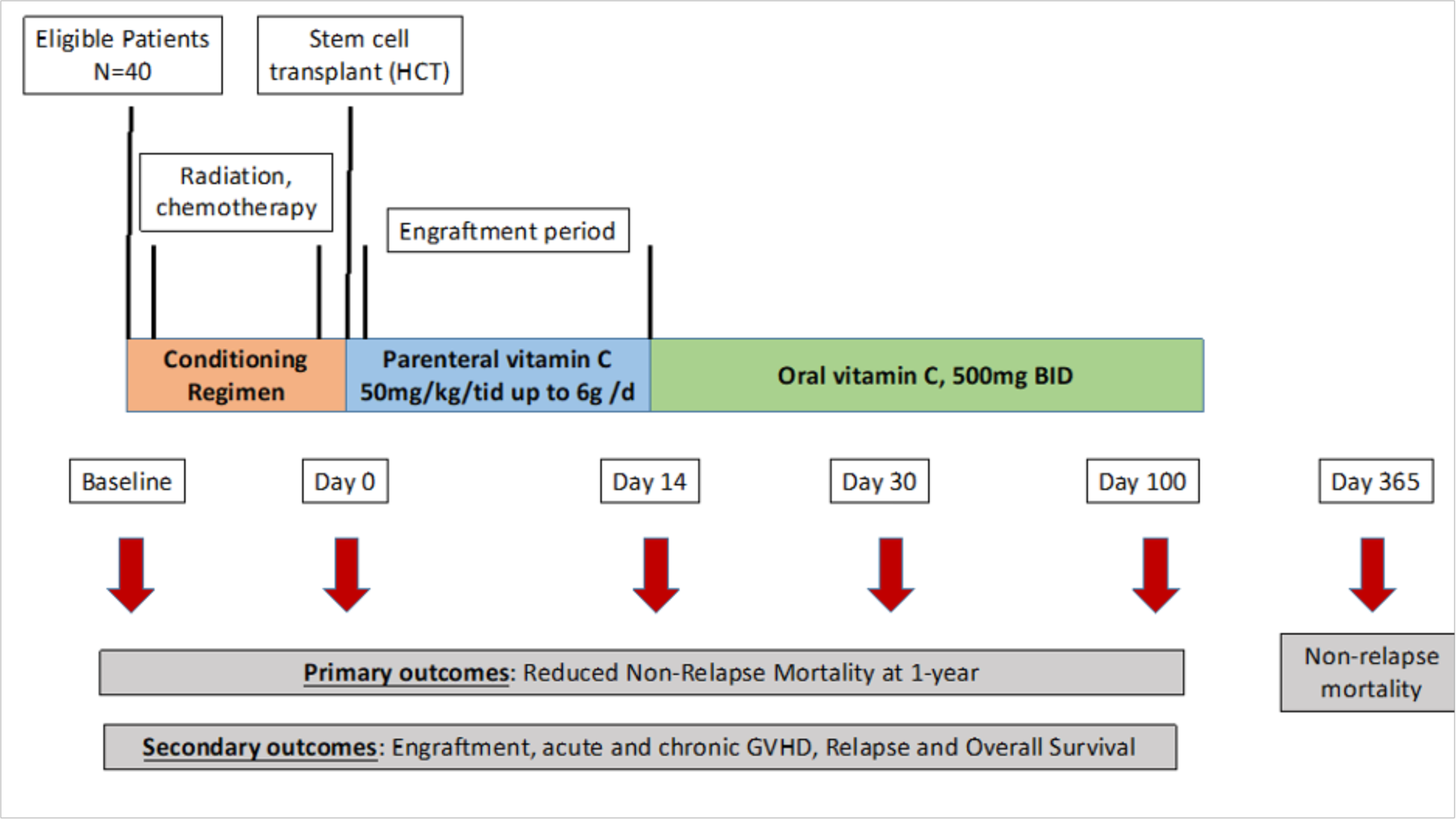
Study schema.

**Supplementary Figure 2.**
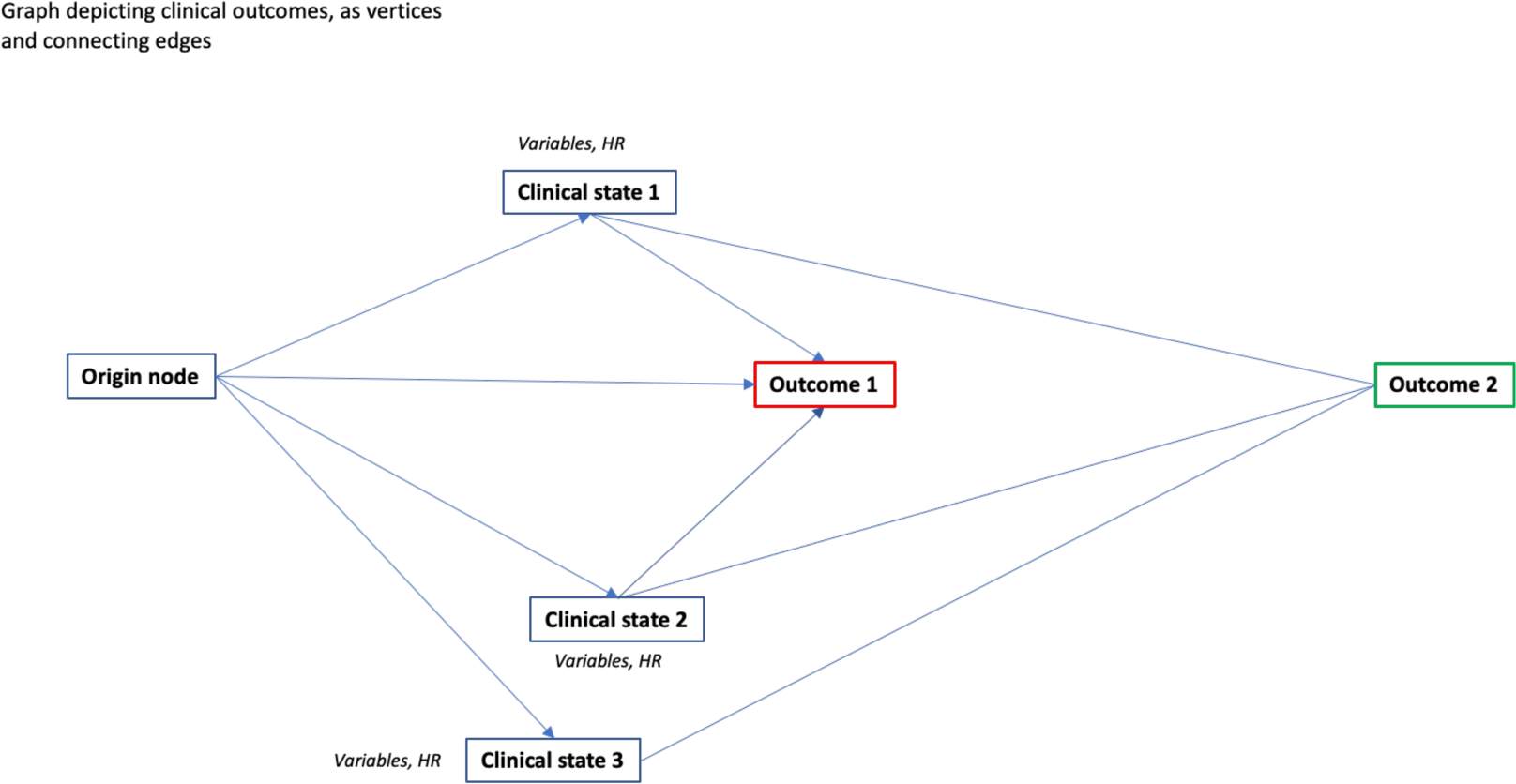
Graph depicting original clinical event, followed by resulting clinical states and outcomes, represented by nodes (vertices) and transition between these nodes represented by edges. The clinical state nodes are first tier transitional states, which lead to the eventual clinical outcome represented by second tier nodes. Edges can be unidirectional or bidirectional, and path along the graph will be determined by prognostic variables. Other intermediary steps may be added as more information becomes available.

**Supplementary Figure 3.**
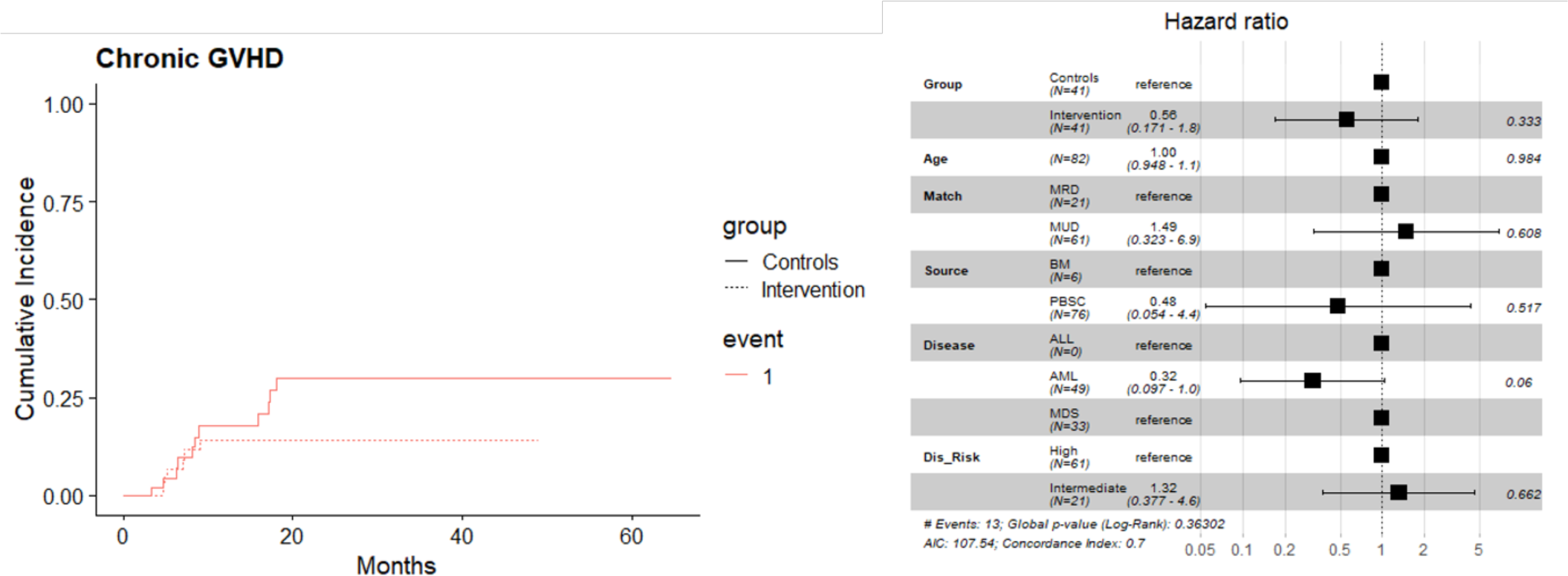
(A) CI curves depicting lower severe cGVHD in the VC treated patients. (B) In a multivariate model this was significant (HR = 0.15, 95% CI: 0.03, 0.71, p-value = 0.017).

**Supplementary Figure 4.**
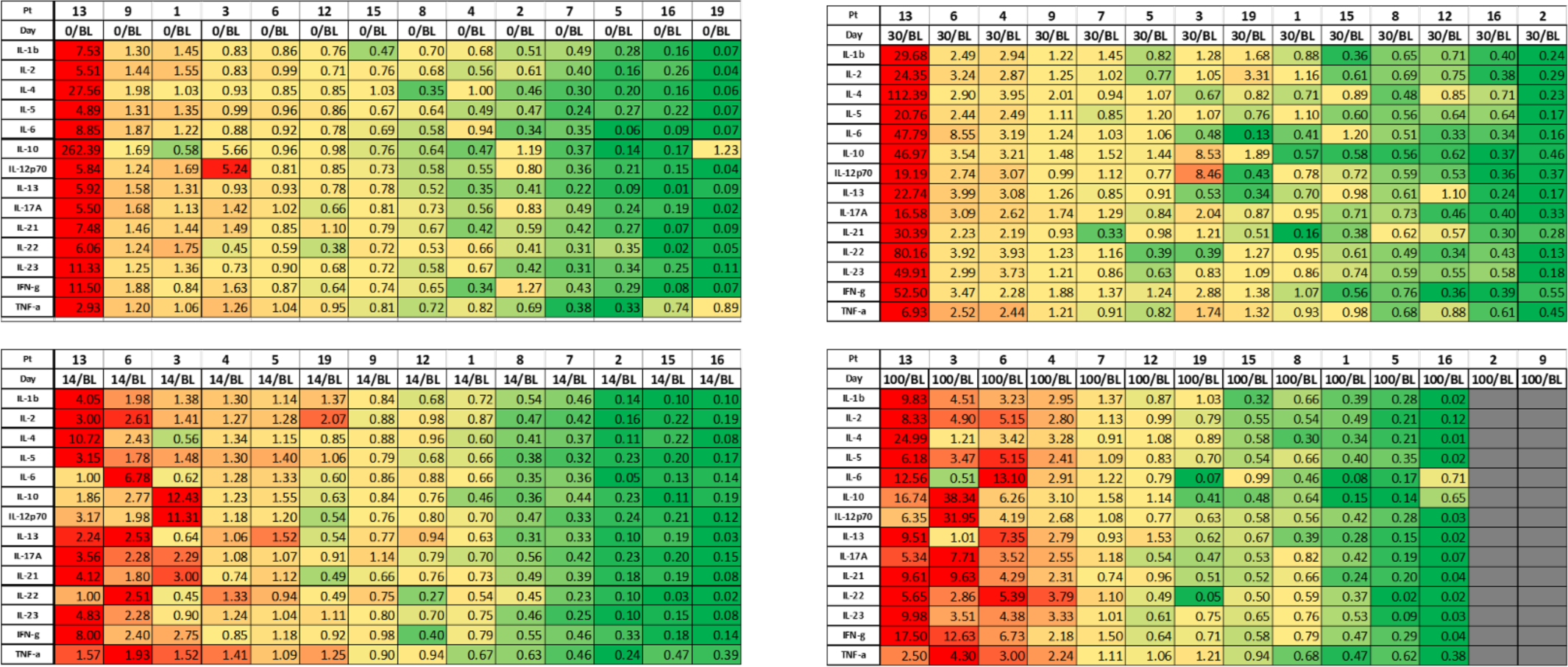
Ratio of cytokine levels at specific time points (days) compared to baseline; day0 /baseline, day 14/baseline, day 30/baseline & day100/baseline. Green signifies a reduction in cytokine levels, while red signifies an increase. Each column gives data for unique patients.

**Supplementary Figure 5.**
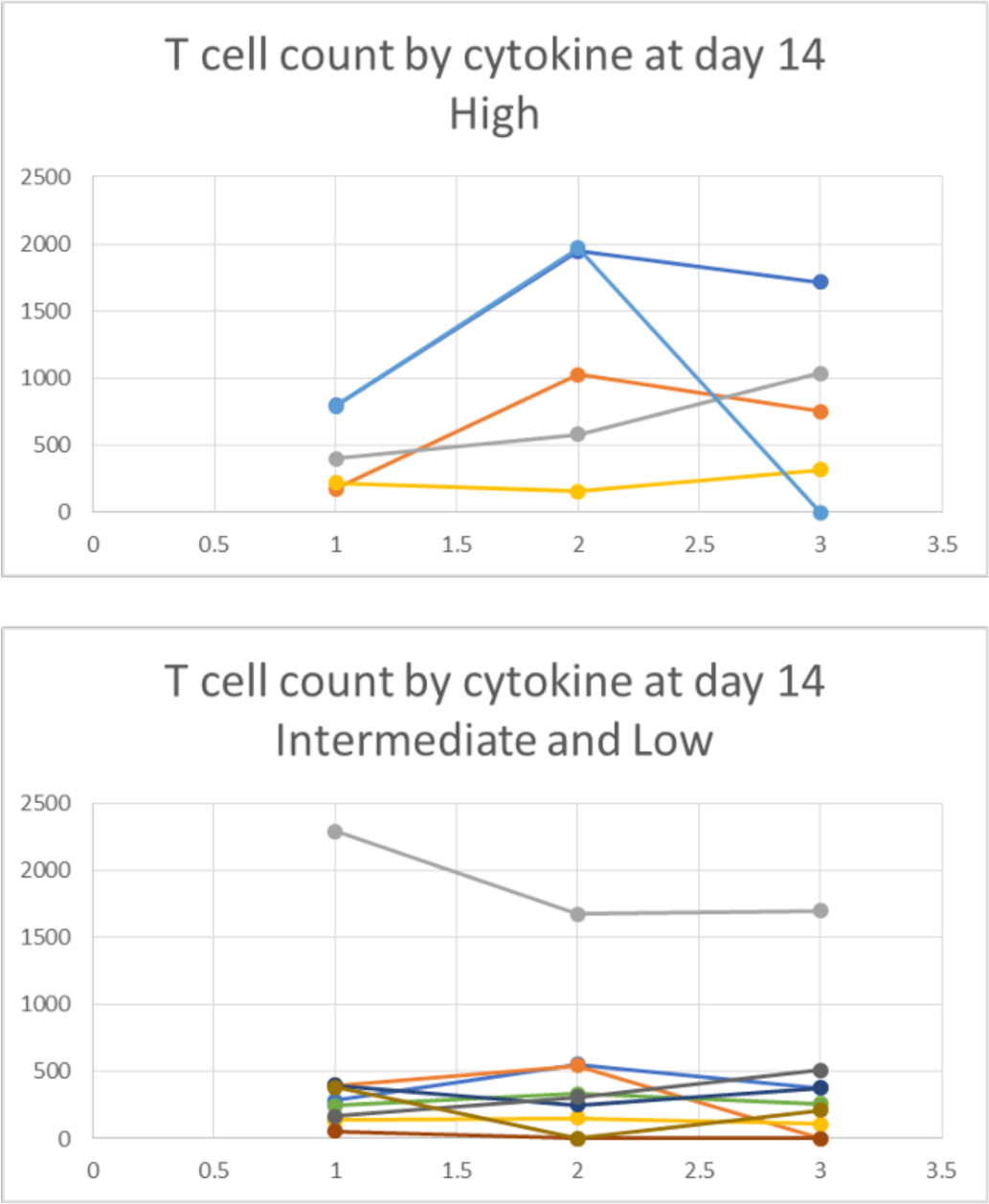
T cell count by cytokine group on day 14. High-13, 6, 3, 4 & 5; Intermediate 19, 9, 12 & 1; Low 8, 7, 2, 15 & 16. X-axis, months post-transplant; Y axis, T cell count/μL.

**Supplementary Table 1:** CTCAE Version 5 Toxicities.

| Body System | Adverse Event | Toxicity Grades (NCI Common Terminology Criteria for Adverse Events V5) |  |  |  |
| --- | --- | --- | --- | --- | --- |
|  |  | 3 | 4 | 5 | TOTAL |
| Blood and lymphatic system disorders | Febrile neutropenia | 4 |  |  | 4 |
| Cardiac disorders | Heart failure | 1 |  |  | 1 |
|  | Sinus tachycardia | 1 |  |  | 1 |
|  | Ventricular tachycardia | 2 |  |  | 2 |
| Ear and labyrinth disorders | Hearing impaired | 1 |  |  | 1 |
| Eye disorders | Optic nerve disorder | 1 |  |  | 1 |
|  | Vision decreased | 1 |  |  | 1 |
| Gastrointestinal disorders | Diarrhea | 9 |  |  | 9 |
|  | Nausea | 5 |  |  | 5 |
|  | Mucositis oral | 10 | 1 |  | 11 |
|  | Small intestinal mucositis | 8 |  |  | 8 |
| General disorders and administration site conditions | Fatigue | 1 |  |  | 1 |
|  | Fever | 1 |  |  | 1 |
| Hepatobiliary disorders | Sinusoidal obstruction syndrome | 2 |  |  | 2 |
| Infections and infestations | Appendicitis | 1 |  |  | 1 |
|  | Epstein-Barr virus infection reactivation | 8 |  |  | 6 |
|  | Cytomegalovirus infection reactivation | 11 |  |  | 9 |
|  | Enterocolitis infectious | 1 |  |  | 1 |
|  | Fungemia | 1 |  |  | 1 |
|  | Herpes simplex reactivation | 1 |  |  | 1 |
|  | Lung infection | 6 |  |  | 6 |
|  | Sepsis | 6 |  | 5 | 11 |
|  | Skin infection | 1 |  |  | 1 |
|  | Urinary tract infection | 7 |  |  | 7 |
|  | Viremia | 3 |  |  | 3 |
| Injury, poisoning and procedural complications | Fracture | 2 |  |  | 2 |
| Investigations | Blood bilirubin increased | 1 |  |  | 1 |
|  | Anorexia | 4 |  |  | 4 |
| Metabolism and nutrition disorders | Hyperglycemia | 4 |  |  | 4 |
|  | Hypernatremia | 1 |  |  | 1 |
|  | Hypoglycemia | 1 |  |  | 1 |
|  | Tumor lysis syndrome | 1 |  |  | 1 |
| Musculoskeletal and connective tissue disorders | Back pain | 1 |  |  | 1 |
|  | Generalized muscle weakness | 1 |  |  | 1 |
|  | Muscle weakness lower limb | 1 |  |  | 1 |
| Nervous system disorders | Headache | 4 |  |  | 4 |
|  | Seizure | 1 |  |  | 1 |
|  | Spinal cord compression | 1 |  |  | 1 |
|  | Syncope | 2 |  |  | 2 |
| Psychiatric disorders | Depression | 1 |  |  | 1 |
| Renal and urinary disorders | Acute kidney injury | 1 |  |  | 1 |
| Respiratory, thoracic and mediastinal disorders | Dyspnea | 1 |  |  | 1 |
|  | Hypoxia | 3 |  | 1 | 4 |
| Vascular disorders | Hypotension | 5 |  |  | 5 |
|  | Hypertension | 2 |  |  | 2 |

## Notes

### Competing Interest Statement

The authors have declared no competing interest.

### Clinical Trial

IND #138924, NCT03613727

### Author Declarations

Virginia Commonwealth University Institutional Review Board gave ethical approval for this work.

### Summary of Updates

This manuscript has been revised to add Graph analysis of clinical outcomes. A series of nodes and edges are utilized to depict clinical states and outcomes following transplantation, demonstrating a reduction in GVHD may be responsible for the observed reduction of NRM and improved survival. Figure 1 and 4 have been added.

## References

1 C. Ferry, and Gérard Socié, Busulfan-Cyclophosphamide versus Total Body Irradiation-Cyclophosphamide as Preparative Regimen before Allogeneic Hematopoietic Stem Cell Transplantation for Acute Myeloid Leukemia: What Have We Learned?”. Experimental Hematology 31, 1182–1186 (2003).

2 H. Y. Lai, Teh Ying Chou, Cheng Hwai Tzeng, and Oscar Kuang Sheng Lee., Cytokine Profiles in Various Graft-versus-Host Disease Target Organs Following Hematopoietic Stem Cell Transplantation. Cell Transplantation 21, 2033–2045 (2012).

3 S. e. a. Dietrich, Endothelial Vulnerability and Endothelial Damage Are Associated with Risk of Graft-versus-Host Disease and Response to Steroid Treatment. Biology of Blood and Marrow Transplantation 19, 22–27 (2013).

4 B. L. Scott et al., Myeloablative Versus Reduced-Intensity Hematopoietic Cell Transplantation for Acute Myeloid Leukemia and Myelodysplastic Syndromes. J Clin Oncol 35, 1154–1161 (2017).

5 Chae, Y. S. et al. 2007. “New Myeloablative Conditioning Regimen with Fludarabine and Busulfan for Allogeneic Stem Cell Transplantation: Comparison with BuCy2.” Bone Marrow Transplantation 40(6): 541–47.

6 N. Molina, Ana Carolina Morandi, Anaysa Paola Bolin, and Rosemari Otton., Comparative Effect of Fucoxanthin and Vitamin C on Oxidative and Functional Parameters of Human Lymphocytes. International Immunopharmacology 22 (2014).

7 J. L. e. a. Kuck, Ascorbic Acid Attenuates Endothelial Permeability Triggered by Cell-Free Hemoglobin. Biochemical and Biophysical Research Communications 495, 433–437 (2018).

8 A. A. e. a. Fowler, Effect of Vitamin C Infusion on Organ Failure and Biomarkers of Inflammation and Vascular Injury in Patients with Sepsis and Severe Acute Respiratory Failure: The CITRIS-ALI Randomized Clinical Trial. Journal of the American Medical Association 322, 1261–1270 (2019).

9 Y. Nannya, Akihito Shinohara, Motoshi Ichikawa, and Mineo Kurokawa., Serial Profile of Vitamins and Trace Elements during the Acute Phase of Allogeneic Stem Cell Transplantation. Biology of Blood and Marrow Transplantation 20, 430–434 (2014).

10 G. B. Dahl, R. I. Jeppsson, and H. J. Tengborn, Vitamin Stability in a Tpn Mixture Stored in an Eva Plastic Bag. Journal of Clinical Pharmacy and Therapeutics 11, 271–279 (1986)

11 M. e. a. Rasheed, Reduced Plasma Ascorbic Acid Levels in Recipients of Myeloablative Conditioning and Hematopoietic Cell Transplantation.” European Journal of Haematology European Journal of Haematology 103, 329–334 (2019).

12 Ferrara JLM, Chaudhry MS. GVHD: biology matters. Blood Adv. 2018 Nov 27;2(22):3411–3417.

13 Teshima T, Reddy P, Zeiser R. Acute Graft-versus-Host Disease: Novel Biological Insights. Biol Blood Marrow Transplant. 2016 Jan;22(1):11–6.

14 Piper C, Drobyski WR. Inflammatory Cytokine Networks in Gastrointestinal Tract Graft vs. Host Disease. Front Immunol. 2019 Feb 22;10:163.

15 Wang W, Hu L, Chang S, Ma L, Li X, Yang Z, Du C, Qu X, Zhang C, Wang S. Total body irradiation-induced colon damage is prevented by nitrate-mediated suppression of oxidative stress and homeostasis of the gut microbiome. Nitric Oxide. 2020 Sep 1;102:1–11.

16 Hu L, Yin X, Zhang Y, Pang A, Xie X, Yang S, Zhu C, Li Y, Zhang B, Huang Y, Tian Y, Wang M, Cao W, Chen S, Zheng Y, Ma S, Dong F, Hao S, Feng S, Ru Y, Cheng H, Jiang E, Cheng T. Radiation-induced bystander effects impair transplanted human hematopoietic stem cells via oxidative DNA damage. Blood. 2021 Jun 17;137(24):3339–3350.

17 Chevion S, Or R, Berry EM. The antioxidant status of patients subjected to total body irradiation. Biochem Mol Biol Int. 1999 Jun;47(6):1019–27.

18 Umegaki K, Sugisawa A, Shin SJ, Yamada K, Sano M. Different onsets of oxidative damage to DNA and lipids in bone marrow and liver in rats given total body irradiation. Free Radic Biol Med. 2001 Nov 1;31(9):1066–74.

19 Saga R, Uchida T, Takino Y, Kondo Y, Kobayashi H, Kinoshita M, Saitoh D, Ishigami A, Makishima M. Radiation-induced gastrointestinal syndrome is exacerbated in vitamin C-insufficient SMP30/GNL knockout mice. Nutrition. 2021 Jan;81:110931.

20 Fujii J, Osaki T, Bo T. Ascorbate Is a Primary Antioxidant in Mammals. Molecules. 2022 Sep 21;27(19):6187.

21 Kuck JL, Bastarache JA, Shaver CM, Fessel JP, Dikalov SI, May JM, Ware LB. Ascorbic acid attenuates endothelial permeability triggered by cell-free hemoglobin. Biochem Biophys Res Commun. 2018 Jan 1;495(1):433–437.

22 Zelikson V, Simmons G, Raman N, Krieger E, Rebiero A, Hawks K, Aziz M, Roberts C, Chesney A, Reed J, Gress R, Toor A. Dynamical Systems Modeling of Early-Term Immune Reconstitution with Different Antithymocyte Globulin Administration Schedules in Allogeneic Stem Cell Transplantation. Transplant Cell Ther. 2022 Feb;28(2):85.e1–85.e9.

23 Ottone T, Faraoni I, Fucci G, Divona M, Travaglini S, De Bellis E, Marchesi F, Angelini DF, Palmieri R, Gurnari C, Giansanti M, Nardozza AM, Montesano F, Fabiani E, Lindfors Rossi EL, Cerretti R, Cicconi L, De Bardi M, Catanoso ML, Battistini L, Massoud R, Venditti A, Voso MT. Vitamin C Deficiency in Patients With Acute Myeloid Leukemia. Front Oncol. 2022 Jun 27;12:890344.

24 Cimmino L, Dolgalev I, Wang Y, Yoshimi A, Martin GH, Wang J, Ng V, Xia B, Witkowski MT, Mitchell-Flack M, Grillo I, Bakogianni S, Ndiaye-Lobry D, Martín MT, Guillamot M, Banh RS, Xu M, Figueroa ME, Dickins RA, Abdel-Wahab O, Park CY, Tsirigos A, Neel BG, Aifantis I. Restoration of TET2 Function Blocks Aberrant Self-Renewal and Leukemia Progression. Cell. 2017 Sep 7;170(6):1079–1095.e20.

25 L. Cimmino, B. G. Neel, I. Aifantis, Vitamin C in Stem Cell Reprogramming and Cancer. Trends Cell Biol 28, 698–708 (2018).

26 Liu J, Min S, Kim D, Park J, Park E, Pei S, Koh Y, Shin DY, Byun JM, Ko M, Yoon SS, Hong J. Pharmacological GLUT3 salvage augments the efficacy of vitamin C-induced TET2 restoration in acute myeloid leukemia. Leukemia. 2023 Aug;37(8):1638–1648.

27 Smith-Díaz CC, Magon NJ, McKenzie JL, Hampton MB, Vissers MCM, Das AB. Ascorbate Inhibits Proliferation and Promotes Myeloid Differentiation in *TP53*-Mutant Leukemia. Front Oncol. 2021 Aug 23;11:709543. doi: 10.3389/fonc.2021.709543. PMID: 34497762; PMCID: PMC8419345.

28 Lamontagne F, Masse MH, Menard J, Sprague S, Pinto R, Heyland DK, Cook DJ, Battista MC, Day AG, Guyatt GH, Kanji S, Parke R, McGuinness SP, Tirupakuzhi Vijayaraghavan BK, Annane D, Cohen D, Arabi YM, Bolduc B, Marinoff N, Rochwerg B, Millen T, Meade MO, Hand L, Watpool I, Porteous R, Young PJ, D’Aragon F, Belley-Cote EP, Carbonneau E, Clarke F, Maslove DM, Hunt M, Chassé M, Lebrasseur M, Lauzier F, Mehta S, Quiroz-Martinez H, Rewa OG, Charbonney E, Seely AJE, Kutsogiannis DJ, LeBlanc R, Mekontso-Dessap A, Mele TS, Turgeon AF, Wood G, Kohli SS, Shahin J, Twardowski P, Adhikari NKJ; LOVIT Investigators and the Canadian Critical Care Trials Group. Intravenous Vitamin C in Adults with Sepsis in the Intensive Care Unit. N Engl J Med. 2022 Jun 23;386(25):2387–2398.

